# Adoption of the OMOP CDM for Cancer Research using Real-world Data: Current Status and Opportunities

**DOI:** 10.1101/2024.08.23.24311950

**Authors:** Liwei Wang, Andrew Wen, Sunyang Fu, Xiaoyang Ruan, Ming Huang, Rui Li, Qiuhao Lu, Andrew E Williams, Hongfang Liu

**Affiliations:** McWilliams School of Biomedical Informatics, The University of Texas Health Science Center at Houston, TX, USA; Clinical and Translational Science Institute Tufts Medical Center Boston US; Institute for Clinical Research and Health Policy Studies Tufts Medical Center Boston US

**Keywords:** Real-world data, cancer research, Observational Health Data Sciences and Informatics (OHDSI) network, Observational Medical Outcomes Partnership (OMOP), Common Data Model (CDM), scoping review

## Abstract

**Background:** The Observational Medical Outcomes Partnership (OMOP) common data model (CDM) that is developed and maintained by the Observational Health Data Sciences and Informatics (OHDSI) community supports large scale cancer research by enabling distributed network analysis. As the number of studies using the OMOP CDM for cancer research increases, there is a growing need for an overview of the scope of cancer research that relies on the OMOP CDM ecosystem.

**Objectives:** In this study, we present a comprehensive review of the adoption of the OMOP CDM for cancer research and offer some insights on opportunities in leveraging the OMOP CDM ecosystem for advancing cancer research.

**Materials and Methods:** Published literature databases were searched to retrieve OMOP CDM and cancer-related English language articles published between January 2010 and December 2023. A charting form was developed for two main themes, i.e., clinically focused data analysis studies and infrastructure development studies in the cancer domain.

**Results:** In total, 50 unique articles were included, with 30 for the data analysis theme and 23 for the infrastructure theme, with 3 articles belonging to both themes. The topics covered by the existing body of research was depicted.

**Conclusion:** Through depicting the status quo of research efforts to improve or leverage the potential of the OMOP CDM ecosystem for advancing cancer research, we identify challenges and opportunities surrounding data analysis and infrastructure including data quality, advanced analytics methodology adoption, in-depth phenotypic data inclusion through NLP, and multisite evaluation.

## INTRODUCTION

Throughout the 21^st^ century, cancer has been a major cause of premature death internationally^1^, leading to substantial research interest. A promising avenue by which can be studied is via observational research, which holds great promise for generating real-world evidence and unique insights, e.g., into patients, treatments, and outcomes.^2,3^ This avenue significantly contributes to advancing clinical knowledge and shaping medical practices.^4^ The primary sources of observational health data encompass electronic health records (EHRs), insurance/administrative claims, hospital billing, clinical registries, and longitudinal surveys.^5^ Given the promise shown in observational research, maximizing the potential of such data is crucial for effective cancer studies, high-quality cancer care, and improved cancer care management.

In particular, conducting multicenter studies is a common strategy used in observational clinical research that allows for improved generalizability of the results, and consequently, improved efficiency. To promote multicenter observational studies, distributed research networks have emerged in recent years, such as the Observational Health Data Sciences and Informatics (OHDSI),^6^ the Agency for Healthcare Research and Quality (AHRQ)-supported projects,^7^ the National Patient-Centered Clinical Research Network (PCORnet)^8^ and the Electronic Medical Records and Genomics (eMERGE) network.^9^ Among these efforts, OHDSI supplies both a common data model (CDM) and the concept representation (terminology) for standardization to support federated analytics, showing great potential for large-scale observational cancer studies.^10,11^ The OHDSI network adopts the CDM developed as part of the Observational Medical Outcomes Partnership (OMOP) to represent data from disparate sources in a standardized format through data normalization processes. A key benefit of such a network-based federated approach is that data holders can maintain their patient-level databases locally, allowing for collaboration through the distributed research network on systematic analytics, increased sample size, heterogeneous patient populations that are geographically dispersed and racially and ethnically diverse, enhanced research generalizability and reproducibility while still maintaining patient confidentiality.

Two previous related reviews have been done on the OMOP CDM. One focused on the adoption of the OMOP CDM in the field of observational patient data research, which delineated the trend over a 5-year period between 2016 and early 2021 by analyzing metadata and topics of literature. ^12^ Results confirmed the increasing importance of the OMOP CDM in conducting network studies internationally within the medical domain. Following that, another review investigated the potential applicability of the OMOP CDM in cancer prediction and how comprehensively the genomic vocabulary extension of the OMOP CDM can serve the needs of AI-based predictions based on the literature between 2016 and 2021.^13^ This study found that the OMOP CDM serves as a solid base to enable a decentralized use of AI in early prediction, diagnosis, personalized cancer treatment, and in discovering important biological markers. While these studies have established the potential for the OMOP CDM for cancer research, the scope of the adoption of the OMOP CDM for cancer research is not well understood. This paper aims to bridge this gap by presenting a comprehensive outline for researchers in the field of cancer study leveraging the OMOP CDM, and guide them to several unexplored research gaps.

## METHODS

Given our objective to explore the scope of the OMOP CDM for cancer studies, we opted for a scoping review. Scoping reviews have been described as an ideal tool for assessing the breadth and extent of a body of literature on a given topic, offering a comprehensive overview of its primary focus and coverage.^14^ We conducted this scoping review with the following five stages based on the framework from Arksey and O’Malley,^15^ and the Preferred Reporting Items for Systematic Reviews and Meta-Analyses extension for scoping reviews.^16^

### Identifying the Research Question

In this scoping literature review, we aimed to identify 1) the extent of cancer data analysis utilizing the OHDSI/OMOP CDM, 2) the maturity of OHDSI/OMOP CDM as an ecosystem infrastructure for cancer research, and 3) challenges and opportunities from the above two themes for potential future investigations.

### Identifying Relevant Studies

We included articles published from January 1, 2010 to December 31, 2023. Only studies written in English were considered. Literature databases surveyed included Journals@Ovid@TMC Library (subscribed full text), Journals@Ovid (some full text), Ovid MEDLINE(R) and Epub Ahead of Print, In-Process, In-Data-Review & Other Non-Indexed Citations and Daily <1946 to January 12, 2024>; IEEE Xplore; PubMed; Web of Science and Embase. A detailed description of the search strategies for articles using OHDSI OMOP for cancer related studies is provided in ***Appendix 1***.

### Study Selection

All the titles and abstracts after deduplication were screened, and the publications were included if OHDSI/OMOP CDM was used for cancer related studies. We excluded publications if they were

1. Not a full-text paper
2. Retrieved by irrelevant term matching
3. Not using OHDSI/OMOP CDM
4. Not cancer focused
5. Not a research paper
6. Not written in English

A second round of full-text screening was done to ensure all publications met the inclusion and exclusion criteria.

### Charting the Relevant Studies

Standardized charting templates were created to summarize pertinent publications. The information of interest was organized around two main themes: data analysis and infrastructure. Two reviewers were assigned to each article, and tasked with independently extracting the information. Consensus was achieved after discussing disagreements between the two reviewers or consultation with a third reviewer.

Shared data elements extracted from the two themes include publication year, data sources, geographic region, and cancer type.

The data analysis theme includes both observational studies and data mining studies. We developed our data extraction schema partially based on the STROBE (strengthening the reporting of observational studies in epidemiology) checklist^17^, a reporting guideline that describes core considerations for observational research. Data elements to extract include objectives, geographic region, cohort size, target domain (disease, drug, etc.), analysis type (SQL, machine learning, statistical analysis, etc.), OHDSI tool used, study period, study design (cohort, case-control, and cross-sectional studies for observational study, ML, or phenotyping, etc.), risk factors explored if applicable, variables (diagnosis, procedures, etc. based on the OHDSI CDM table names), statistical methods, NLP usage, number of datasets. To facilitate subsequent analysis, we aggregated variables based on the OHDSI CDM table names (https://ohdsi.github.io/CommonDataModel/).

A data extraction schema for the infrastructure theme was developed to encompass key components including source data warehouse type (local EHR, claims data, etc.), source data type (diagnoses, procedures, etc. based on the OHDSI CDM table names), mapping coverage, main challenges in ETL, evaluation method of mapping, data model extension, limitation of data model (data element not specified, no definition, etc.) and entity linking/normalization method.

### Collating, Summarizing, and Reporting the Results

The results obtained from the data charting for each theme were summarized, analyzed, and visualized to present an overview of the application of OHDSI/CDM in the field of cancer.

## RESULTS

The article selection process is shown in Figure 1. After identifying the included articles, the study team performed a comprehensive full-text review of the resulting 50 studies. There are 30 studies focusing on data analysis and 23 on data infrastructure. Among them, 3 articles (published in 2018, 2020, and 2021) belong to both data analysis and infrastructure. articles (charting items) are provided in the ***Data Supplement***.

**Figure 1.**
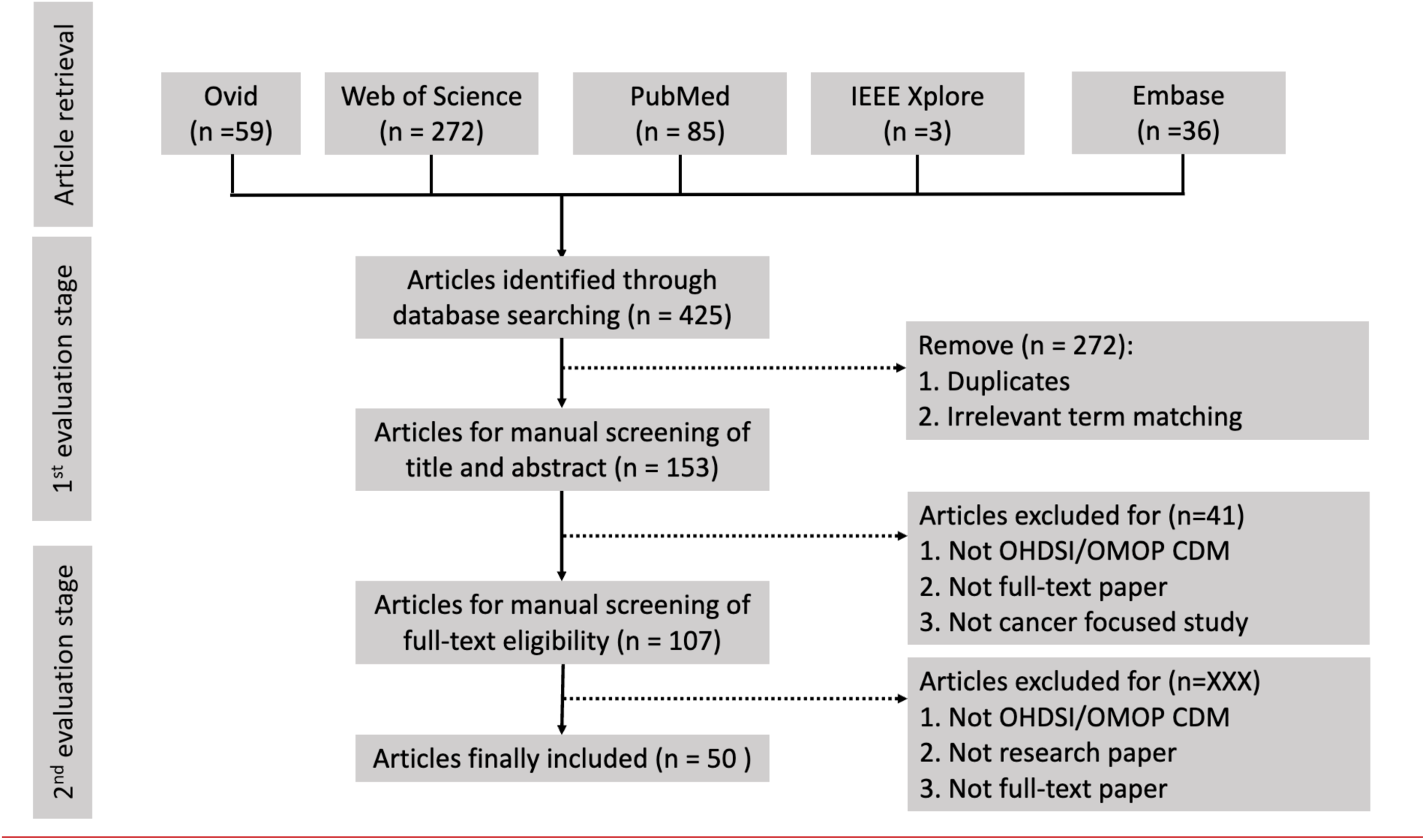
Article selection process.

All extracted data from the

### Overview analysis

Figure 2 shows the distribution of studies stratified by the two themes, i.e., Infrastructure and Data analysis, with 3 articles (published in 2018, 2020, 2021) belonging to both data analysis and infrastructure. Though we collected articles from 2010, the first article included in our study was published in 2017, and data analysis papers showed an increasing trend from 2018 to 2022 (Figure 2A).

**Figure 2.**
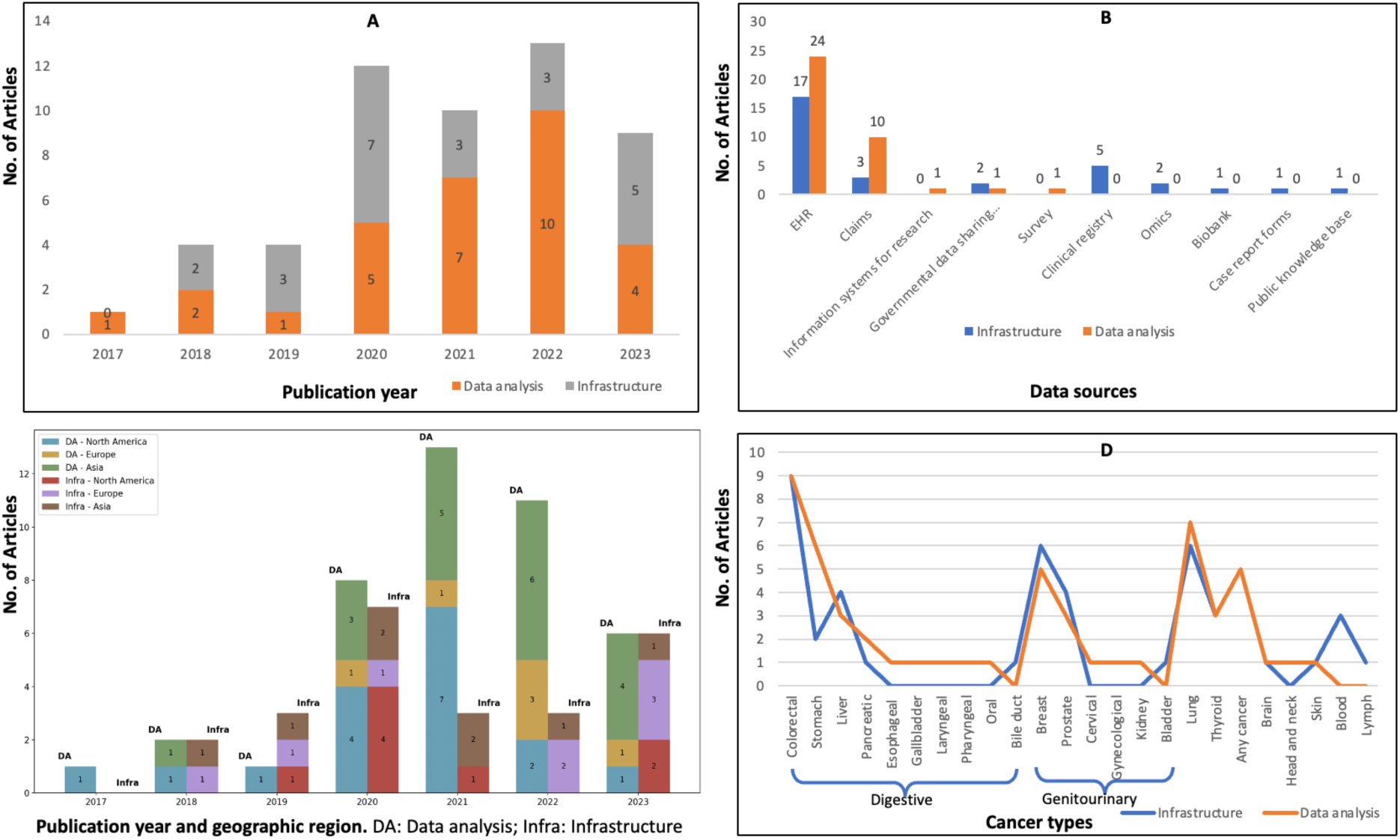
A: Distribution of all articles across publication year (A), data sources (B), publication year and geographic region (C) and cancer types (D), stratified by data analysis and infrastructure.

Figure 2B compares the data sources used between Infrastructure and Data Analysis studies. One article may include more than one data source. In general, usage of EHR data has been the mainstream in both themes, with claims data being another important source for the data analysis theme. EHR was used in combination with one additional data source in 7 infrastructure-themed articles (claims and survey),^19,21–26^ and 7 data analysis-themed articles (claims, registry, and omics).^10,19,27–31^ EHR was used with two additional data sources (claims and registry) in only 1 infrastructure-themed article. ^32^ Compared with the data analysis theme, several new types of data sources emerged for infrastructure construction, including clinical registries, omics, Biobank, case report forms, and public knowledge bases. Table 1 lists the references of data sources in each theme.

**Table 1:** Comparison of Infrastructure and Data analysis in data sources.

| Data sources | Infrastructure (n=23) | Data analysis (n=30) |
| --- | --- | --- |
| EHR | 17<sup>18-26,32-39</sup> (74%) | 24<sup>10,11,18-20,27-31,40-53</sup> (80%) |
| Claims | 3<sup>19,22,32</sup> (13%) | 10<sup>10,19,27-29,31,54-57</sup> (33.3%) |
| Information systems for research | 0 | 1<sup>58</sup> (3.3%) |
| Governmental data sharing | 2<sup>24,59</sup> (8.7%) | 1<sup>60</sup> (3.3%) |
| Survey | 0 | 1<sup>30</sup> (3.3%) |
| Clinical registry | 5<sup>21,25,32,61,62</sup> (21.7%) | 0 |
| Omics | 2<sup>23,26</sup> (8.7%) | 0 |
| Biobank | 1<sup>63</sup> (4.3%) | 0 |
| Case report forms | 1<sup>64</sup> (4.3%) | 0 |
| Public knowledge base | 1<sup>65</sup> (4.3%) | 0 |

Figure 2C shows the distribution of papers geographically in the North America, Asia and Europe. The USA, South Korea, and Germany stood out as the leading countries in each geographic region in the infrastructure and data analysis themes. More detailed analyses are shown in the results of the infrastructure and data analysis themes below. Figure 2D shows a similar trend of cancer types between the data analysis and the infrastructure theme, with a spike in the infrastructure theme highlighting the need for data construction for blood and lymph. The cancer types studied in the two themes covered a broad range, and the variation in the number of articles focused on each type was present. Table 2 lists the references (n>1) of the specific cancer types in each theme.

**Table 2:** Comparison of Infrastructure and Data analysis in cancer types.

| Classification | Specific cancer | Infrastructure | Data analysis |
| --- | --- | --- | --- |
| Digestive system | Colorectal | 9<sup>18,19,22,25,37-39,59,64</sup> | 9<sup>18,19,43,44,50-52,54,57</sup> |
|  | Stomach | 2<sup>22,25</sup> | 6<sup>43,44,54-56,58</sup> |
|  | Liver | 4<sup>22,25,64,66</sup> | 3<sup>43,44,54</sup> |
|  | Pancreatic | 1<sup>25</sup> | 2<sup>43,54</sup> |
| Genitourinary system | Breast | 6<sup>19,22,34,39,62,64</sup> | 5<sup>19,43,44,48,54</sup> |
|  | Prostate | 4<sup>22,33,35,64</sup> | 3<sup>43,53,54</sup> |
| Respiratory system | Lung | 6<sup>19,22,26,35,62,64</sup> | 7<sup>19,30,40,43,44,49,54</sup> |
| Endocrine system | Thyroid | 3<sup>22,36,37</sup> | 3<sup>41,45,54</sup> |
| Blood and blood forming organs | Blood | 3<sup>21,34,63</sup> | 0 |

Figure 3 compared the clusters based on cancer types and CDM table names (variables) between the infrastructure and Data analysis themes. Compared with the data analysis theme (Figure 3B), richer variable tables were involved in the infrastructure theme (Figure 3A) for colorectal cancer including Episode, Episode_event, Fact_relationship, Location, Note, Note_NLP, Specimen, Care_site, Unique_conditions, Uniqe_observations and Unique_procedures; for breast cancer including Device_exposure, lymphovascular cancer including vascular Note, thyroid cancer including Note, and blood cancer including specimen.

**Figure 3.**
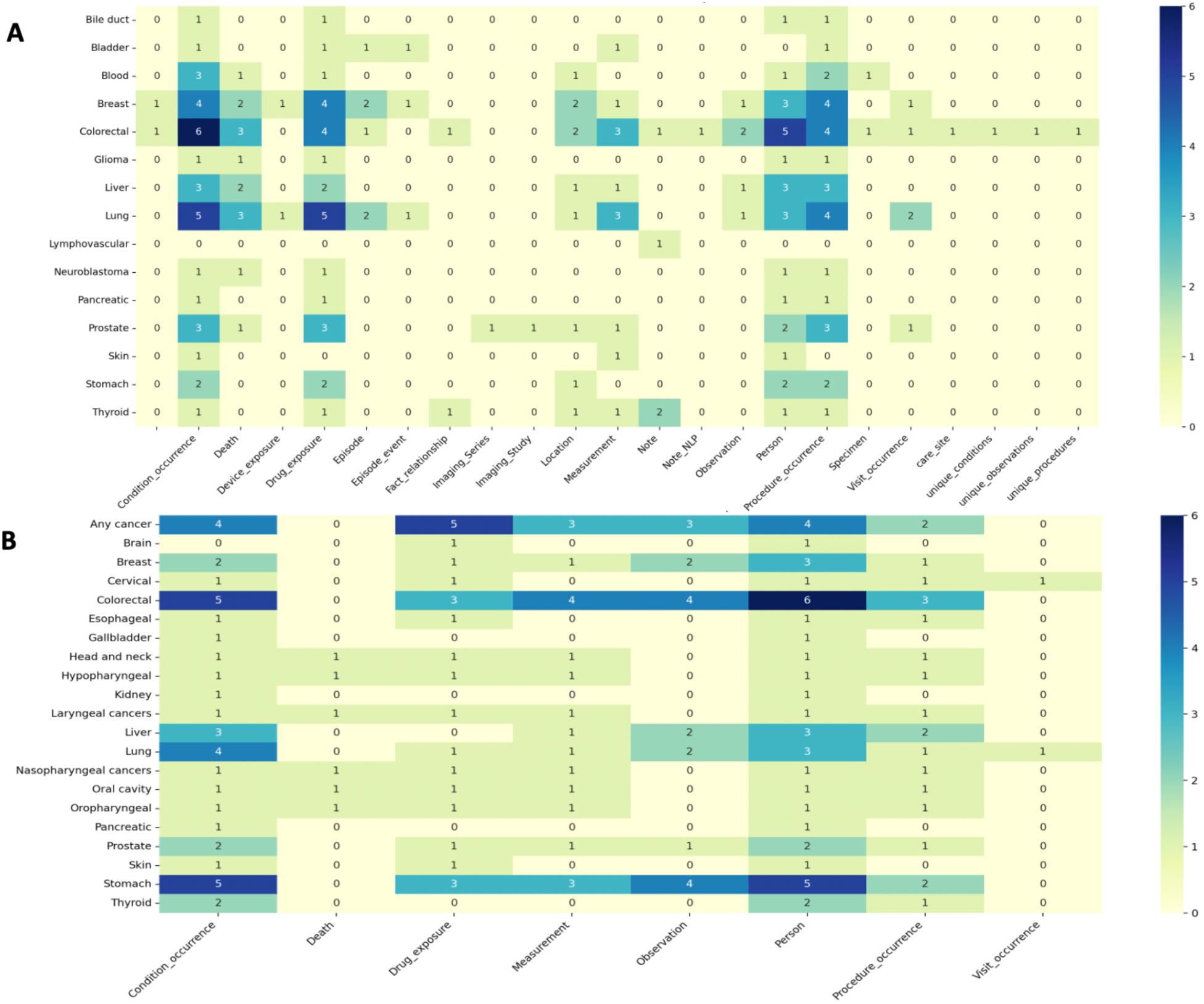
Comparison of clusters based on article numbers of co-occurrence of cancer types and CDM tables (variables). A: Infrastructure theme, B: Data analysis theme.

### Infrastructure theme

Before data analysis can be conducted, the data itself must be present in the OMOP CDM format, and tooling to support that data analysis must exist. In this theme, we will therefore summarize efforts to develop reusable tooling and practices to transform data to the OMOP CDM format, as well as expand the OMOP CDM to support additional data, in relation to cancer. A total of 23 studies fell under this category. Broadly speaking, studies done in this category can be divided into 4 subcategories, i.e., infrastructure development, transformation of various source data types to the OMOP CDM, Data Model extensions and development, and Data Linkage and Standardization. Table 3 shows the references of papers in the 4 subcategories.

**Table 3.** A summary of papers in the infrastructure theme.

| Theme | Subcategory | Papers | Papers with some form of evaluation |
| --- | --- | --- | --- |
| Infrastructure | infrastructure development | N=9<sup>18,21-23,25,35,37,39,64</sup> | N=3<sup>21,23,39</sup> |
|  | transformation of various source data types to the OMOP CDM | N=6<sup>19,20,36,38,59,61</sup> | N=3<sup>36,38,61</sup> |
|  | Data Model extensions and development | N=3<sup>26,32,33</sup> | N=2<sup>26,32</sup> |
|  | Data Linkage and Standardization | N=5<sup>24,34,62,63,65</sup> | N=3<sup>34,63,65</sup> |

#### Geographic region and datasets

In terms of geographic region, studies within this category are split equally across three geographic regions with the United States (n=8), ^22,24,25,32,34,63–65^ Europe (n=8), ^21,23,33,39,59,61,62,64^ Asia with South Korea (n=6) ^19,26,35–38^ and China (n=2).^18,66^ Within Europe, Germany is particularly distinct as it participates in 5 of the included studies from that region. ^23,39,59,61,62^

A majority of these articles remain concentrated within a single dataset (n=11), ^23,25,33,35,37,38,61– 63,65,66^ which is reasonable for infrastructure construction efforts. Of the remainder, 4 studies involve 3 datasets, ^19,22,36,39^ 3 studies involve 2 datasets, ^24,26,34^ 1 studies involve 6, ^18^ 1 study involves 8, ^32^ and 1 study involves 20. ^21^ One study did not report a dataset. ^33^

#### OMOP data and model extension

Of the studies (n=8) that sought to extend the OMOP CDM or enrich the data contained within, ^23,26,32,33,36,38,64,65^ 5 sought to extend the model to better support oncology-related data elements, ^32,36,38,64,65^ 2 sought to extend support for –omics data, ^23,26^ and 2 sought to extend support for imaging data. ^33,64^

#### Data mapping and evaluation

A bulk (n=12) ^18,19,22,24,25,33,35,37,59,62,64,66^ of the studies in this category do not report a direct evaluation of the mapping quality into the OMOP CDM. Evaluation metrics were similarly ill-defined, although the most common evaluation was mapping coverage/percentage of source rows that were successfully mapped to the OMOP CDM standard (n=4), ^34,36,38,63^ or the proportion of clinical concepts that could be successfully represented in the OMOP CDM standard (n=2). ^32,61^ Besides the studies reporting evaluation (n=11), two studies ^59,62^ did not have an evaluation of the mapping process but did report a metric of the percentage of concepts that were not representable.

Common themes regarding reported limitations of data mapping include the fact that the OMOP CDM could not represent certain clinically relevant concepts without further extension (n=6) ^23,32,33,59,65,66^ and that some data was not directly available in structured form and required algorithmic normalization (n=3). ^24,38,66^

### Data analysis theme

To better delineate the relationship amongst the various data elements collected, we conducted synthesis analyses for the data analysis theme. Figure 4 shows the linkage between aggregated cancer types, geographic area, study population size, and the study period of the corresponding population. To categorize geographic locations, the global study is defined as a study that includes at least two countries, in contrast to a single-country study, which includes only one country. Global studies (n=6) started from 2020, ^10,18,27,29,31,52^ that accounted for 20% of papers in the data analysis theme. Global collaborations were across North America, Europe, and Asia, including USA, Spain, France, Germany, UK, Denmark, Netherlands, South Korea, and China, with the USA participating in the majority of studies, contributing to 5 out of 6 studies (83.3%). Among the 24 single-country studies, 15 came from South Korea, 6 from the USA, 2 from Denmark and 1 from China.

**Figure 4.**
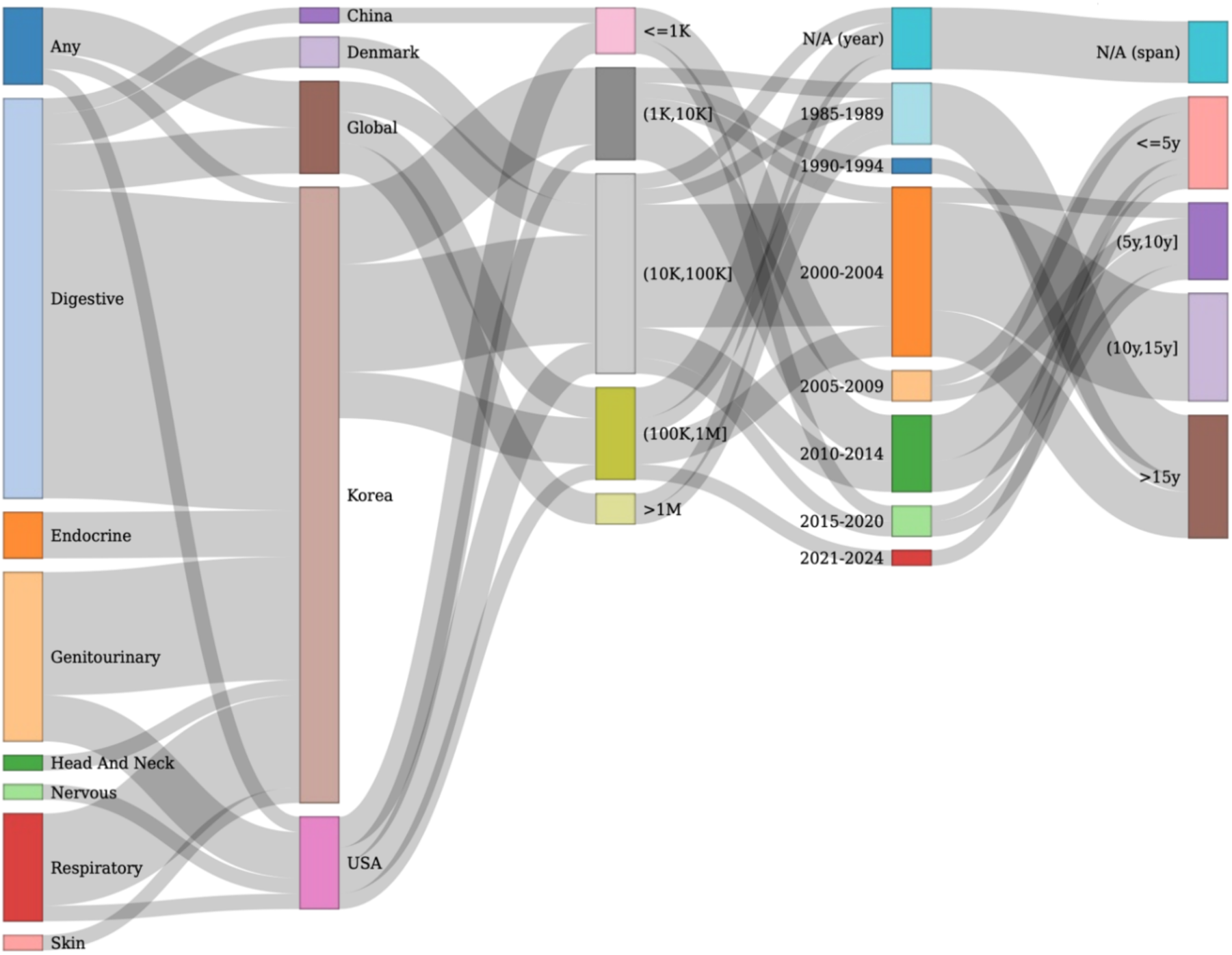
Linkage between the aggregated cancer type, geographic area, cohort size, start year of study, and study period. Analysis based on all countries.

Among 30 studies in the data analysis theme, 15 (50%) studies leveraged multi-site datasets ranging from 2 to 11 individual sites. ^10,11,18,19,27,29,31,40–43,45,47,51,53^ The remaining 15 studies used a single dataset, including 8 from South Korea ^44,46,49,54–58^ 4 studies from USA, ^28,30,48,60^ and 1 each from Denmark, ^50^ China, ^20^ and a collaboration effort between Denmark and Netherland. ^52^ In terms of cancer types and population, 15 studies on the South Korean population covered all cancer types except nervous system (brain cancer), which was exclusively conducted in the US population. ^47^ Six local studies in the USA concentrated on genitourinary, nervous, and respiratory cancers. ^28,30,42,47,48,53^ Denmark ^50,51^ and China ^20^ focused on digestive system cancers in their local studies. While global studies had the capacity to cover more than 1M population, ^29,31^ local studies covered the population ranging from <=1K to 1M. The earliest period started in 1986; two covered the South Korean population, ^19,42^ and one covered the global population. ^49^ Study period of 7 studies exceeded 15 years. ^27,30,31,42,50,51,53^ Four studies didn’t provide the period of the studied population.

As studies from South Korea are disproportionally prevalent compared with other nations, to simplify visualization, Appendix Figure 2 shows the linkage after excluding local studies from South Korea. The upper right of the figure indicates that the study period between 1995 and 1999, and three cancer types, i.e., endocrine, head and neck and skin cancers were dropped out compared with Figure 3A.

Study designs are categorized under two broader groups: “observational study” and “advanced analytics”. The “observational study”, comprising 22 (73.3%) papers, and “advanced analytics” was presented in the relatively minor portion with 8 (26.7%) studies. Table 4 provides a list of references for the study methods.

**Table 4.** References for the study methods.

| Theme | Study methods (n=30) | Subcategories | Papers with some form of evaluation |
| --- | --- | --- | --- |
| Data analysis | Observational study (n=22, 73.3%) | Cohort study | n=16<sup>10,11,27-30,40,42,43,45,46,54-58</sup> |
|  |  | Descriptive study | N=3<sup>19,20,31</sup> |
|  |  | Case control | N=2<sup>44,60</sup> |
|  |  | Cross sectional | N=1<sup>41</sup> |
|  | Advanced analytics (n=8, 26.7%) | Predictive modeling | N=7<sup>18,47,49-53</sup> |
|  |  | Phenotyping | N=1<sup>48</sup> |

Figure 5 illustrates the relationships between target domains, study designs, statistical methods, and variables used across these data analysis studies. The majority (86.7%) of the research efforts focused on two primary domains, i.e., disease (n=14) ^10,18,20,28,30,41,44,47,50–54,60^ and adverse drug events (ADE) (n=12), ^11,27,29,40,42,43,45,46,55–58^ respectively. Other domains included risk factors for emergency department (ED) visits, ^49^ treatment patterns, ^19,31^ and trial eligibility ^48^.

**Figure 5.**
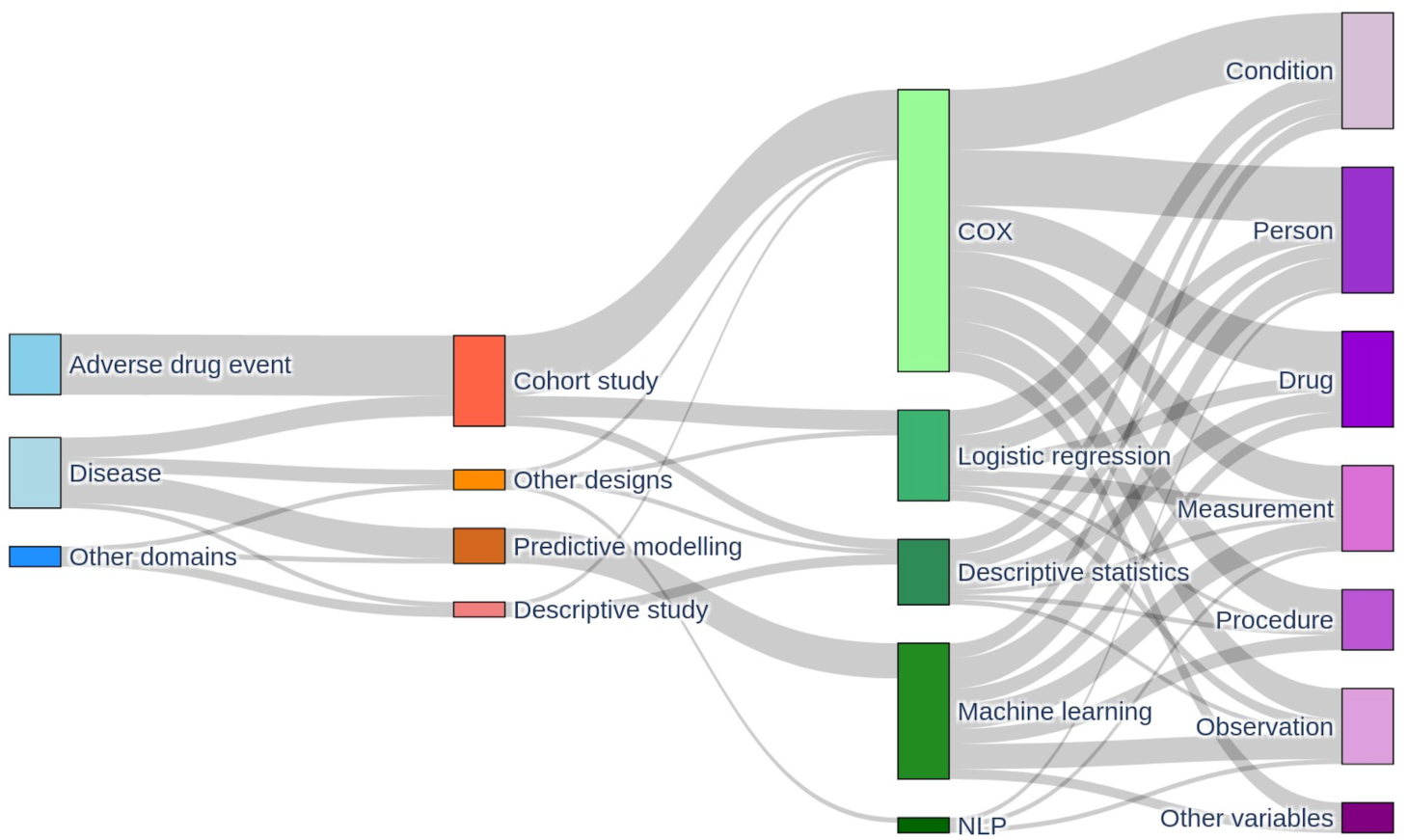
Analysis of target domains, study designs, statistical methods, and variables in the data analysis theme

All the 14 observational studies on ADE exclusively utilized the cohort study design. Conversely, observational studies on diseases include a variety of study designs. Among these, predictive modeling was the dominant approach (n=6), ^18,47,50–53^ while cohort studies ranked second in usage (n=4). ^10,28,30,54^ Specifically, the COX model was the most widely used statistical method in observational studies (n=12), ^11,20,27–29,40,42–45,55,58^ followed by logistic regression (n=5). ^30,41,46,56,57^ Machine learning is the sole method for advanced analytics in predictive modeling study design (n=7). NLP was only employed in an observational study for the trial eligibility via phenotyping.^48^

In data analysis studies, a wide range of variables were leveraged by both statistical and machine learning methods. The most frequently used variables include condition occurrence, person, drug exposure, death, procedure occurrence, measurement, observation, and visit occurrence.

## DISCUSSION

We conducted a scoping review on the adoption of the OMOP CDM for cancer studies since 2010. In the following subsections, we will discuss the extent of cancer data analysis and the maturity of the OMOP CDM as an infrastructural eco-system for cancer research, as well as associated challenges and opportunities for future investigation.

### Status quo

The existence of data analysis-themed studies implies that the data used was prepared sufficiently for the targeted studies while infrastructure-themed studies might imply unmet data management needs. OHDSI was founded in 2008 and started to yield publications in 2010, ^67^ while cancer data analysis studies started in 2017, ^47^ and infrastructure publications started in 2018, global studies started in 2020 ^18,27,31^. Of note, OMOP CDM enabled longitudinal studies spanning 15 years of study period ^27,30,31,42,50,51,53^ and studies with more than 1 million population. ^29,31^ It’s shown that the USA, South Korea, and Germany stood out as the leading countries improving or leveraging OMOP CDM for the cancer domain in each continent, consistent with the previous review study. ^12^ It’s worth noting that most data analytics studies focused on disease and adverse drug events.

### Maturity of the OMOP CDM ecosystem

Examining the cancer types, data sources, and variable CDM tables being studied is helpful in understanding whether real-world data are well-prepared and meet the data needs for downstream analysis. Infrastructure and data analysis showed a roughly consistent trend in the wide range of cancer types they covered. The diverse set of data sources included in the reviewed infrastructure studies suggests that cancer studies require additional data sources beyond the current EHR data-focused ecosystem. Meanwhile, new variable tables, such as Episode, Note_NLP and Specimen, and data model extension for omics and imaging data were involved in the infrastructure theme. It is, therefore, evident that the OMOP CDM ecosystem is still undergoing active development and iteration, which will result in continuous improvement to better support cancer studies.

### Adoption of advanced analytics methodology

Cancer research of the data analysis theme showed a strong preference for observational cohort studies, placing high value on long-term longitudinal analysis for drawing evidence over time. While limited in number, data mining studies were explored to gain predictive insights, suggesting an emerging stream in cancer research within the OMOP CDM framework. Machine learning models were the primary methods, while deep learning and large language model-based approaches remain unexplored. In light of the critical role of data infrastructure, one study presented an overview of the development efforts towards sustainable AI cloud-based platforms for developing, implementing, verifying, and validating trustable, usable, and reliable AI models regarding cancer care provision. ^64^

### In-depth phenotypic data inclusion

It should be noted that a substantial amount of clinically relevant information for cancer is represented in unstructured form. This is particularly the case for information contained within pathology reports, as synoptic reporting is only currently adopted for a minority of cancer types within many institutions. However, limited studies explored NLP methods to build data infrastructure, ^36–38^ and only 1 study leveraged NLP-derived data in the data analysis theme. ^48^ Potential challenges of the current NLP methodology for handling text data were highlighted in these studies, e.g., the limitations of using simple regex in NLP, along with concerns regarding generalizability and systematic evaluation of annotation schemas. ^48,37,38^ We also identified and discussed issues and barriers for wide adoption of cancer NLP in our previous study. ^68^ Despite the challenges, it is critical to incorporate NLP-derived data within OMOP CDM instances for cancer research. In the context of multiple sites and privacy-preserving demands, a federated NLP deployment framework following the RITE-FAIR (Reproducible, Implementable, Transparent, Explainable - Findable, Accessible, Interoperable, and Reusable) principles with scientific rigor and transparent (TRUST) provides a solution towards real-world clinical NLP. ^69,70^

### Data quality

The data quality challenge was primarily related to two sub-types, i.e., accessibility information quality (IQ) and representational IQ. ^71,72^ For accessibility IQ, concerns related to poor record linkage and inaccessible geocoding information were discussed by several studies. ^22,33,34^ Data timeliness was another issue as the current data retrieval and operation process is steward-based and lacks a real-time process (n=2). ^25,39^ Data privacy, security (e.g., data reidentification) and regulatory considerations play a significant role in addressing accessibility IQ. ^25^ Regarding representational IQ, the lack of data standardization, particularly in the context of limitations within OMOP vocabularies, was a continuous challenge. In addition, a substantial portion of the reviewed studies in the infrastructure theme did not perform mapping quality evaluation, which presented a significant gap as variations in such a process can have profound effects on the validity of any downstream use cases. The potential solution for the data standardization and concept mapping problem lies in efforts to derive human-driven consensus amongst multiple use-cases on individual value-sets corresponding to individual clinical entities. Most prolific amongst these efforts is the NLM’s Value Set Authority Center (VSAC) ^73^ which aims to render clinical concept sets publicly available for further reuse and refinement. Beyond that, efforts have been made to create additional tooling allowing for similar functions at an institutional level (with greater human interaction), such as the OHNLP Valueset Workbench. ^74,75^ Nevertheless, each of these tools is relatively standalone and greater effort should be made to integrate similar functionality into current clinical phenotyping workflows.

### Multisite evaluation

Despite the OMOP CDM is designed to support multi-site studies, our review indicates that the majority of studies used single-site data. A lack of multisite evaluation for proposed methods/frameworks,^18,19,22,24,25,33,35,37,59,62,64,66^ and representativeness of research findings due to single site data analysis design. ^28,30,44,46,48,49,54–58,60,50, 52,20^ was shown in the infrastructure and data analysis themes, respectively. Site-specific infrastructural biases within individual data sources further compound these challenges. Overall, the challenges lie in the multifaceted nature of the data ETL and harmonization process, emphasizing the need for comprehensive approaches to overcome technical, regulatory, and operational challenges.

While harmonization of clinical data via the OMOP CDM has vastly improved this state of affairs, the issue persists due to non-standard approaches by which this data is populated, particularly when it comes to concept normalization approaches. This issue is further complicated by the closed nature of many current EHR system licenses, limiting public sharing of developed ETL pipelines and leading to a substantial amount of re-implementation with differing methodologies. In the absence of any change on the EHR license terms, perhaps the best approach is to actively publish concept mappings (e.g., via mechanisms such as the aforementioned Valueset Workbench ^73^) such that they can be reviewed, refined, and re-used later on down the line, particularly in the case of manual mappings and/or NLP-derived mappings from text-based clinical concepts.

## CONCLUSION

In this scoping review, we depicted the status quo of research efforts to improve or leverage the potential of the OMOP CDM ecosystem for advancing cancer research. Our findings revealed that while the OMOP CDM ecosystem has reached a level of maturity that is sufficient to support cancer research, ongoing model development and iteration remains needed to fulfill additional research data needs. Subsequently, we identify challenges and opportunities surrounding data analysis and infrastructure including data quality, advanced analytics methodology adoption, in-depth phenotypic data inclusion through NLP, and multisite evaluation.

## Data Availability

All data produced in the present study are available upon reasonable request to the authors

## ACKNOWLEDGEMENT

This project is supported by the Cancer Prevention Research Institute of Texas (CPRIT) RR230020, National Institute of Aging grant RF1AG072799, National Human Genome Research Institute R01HG12748, and National Library of Medicine R01LM11934.

## AUTHOR CONTRIBUTION

L.W.: conceptualized and designed the study, analyzed the data of the data analysis theme, visualized results and drafted the manuscript; A.W.: conceptualized and designed the study, analyzed the data of the infrastructure theme and drafted the manuscript; F.S.: designed the study, analyzed the data of infrastructural theme and drafted the manuscript; X.R.: designed the study, analyzed the data of the data analysis theme, visualized results and drafted the manuscript; M.H.: analyzed the data of the data analysis theme, visualized results and drafted the manuscript; R.L.: analyzed the data of infrastructural theme and visualized results; Q.L.: analyzed the data of infrastructural theme; A. Wi.: conceptualized, and revised the manuscript; H.L.: conceptualized, supervised, and designed the study and revised the manuscript.

## Appendix

### Summary of studies using NLP

Appendix Table 1 shows the summary of papers using NLP for OMOP-based cancer studies. The data analysis theme has 1 study in the clinical trial domain. ^48^ As a large majority of eligibility criteria is only mentioned in unstructured clinical text besides demographics, NLP was shown to extract eligibility criteria from unstructured clinical notes with high precision and recall. OMOP was used for the computable representation of eligibility criteria. Additionally, we identified 3 papers from the “Infrastructure” theme using South Korean EHRs. ^36–38^ One proposed a framework for hierarchical annotation of textual data and integration into a standardized OMOP-CDM medical database. ^37^ This study utilized topic modeling to identify medical concepts within the unstructured documents and conducted multidimensional validation by identifying associations, such as the association of node positivity with mortality in patients with colorectal cancer. In an effort to transform pathology reports into the CDM, regular expression rules were used to extract clinical and genetic information. ^38^ Manual chart review was conducted for validation but no result was reported for the NLP performance. In another study, thyroid cancer diagnosis and cancer stage information were extracted from pathology reports and whole-body scan reports. ^36^ Of the 4 studies using NLP, 3 used rule-based methods, only 1 study worked on multi-site data from three metropolitan university hospitals, ^36^ and 3 conducted NLP evaluation or validation in only one institution. ^48,37,38^

**Appendix Table 1.** A summary of papers adopting NLP.

| Themes | Paper | Publication year | NLP methods | Document type | Extracted data elements | Geographic area |
| --- | --- | --- | --- | --- | --- | --- |
| Advanced analysis | Automatic trial eligibility surveillance .... | 2019 | rule-based and ML-based | clinical notes | ECOG, cancer staging, biomarkers, functional status and menopausal status | U.S. |
| Infrastructure | Transformation of Pathology Reports Into ... | 2020 | rule-based | Pathology Reports | Cancer diagnosis, genetics | South Korea |
|  | A Framework (SOCRA<sub>TE</sub>x) for Hierarchical Annotation ... | 2021 | topic modeling manual annotation | Pathology reports, radiology reports, and admission notes | Not pre-defined | South Korea |
|  | Transforming Thyroid Cancer Diagnosis and Staging Information ... | 2022 | rule-based | pathology reports and whole-body scan reports | Cancer diagnosis and staging | South Korea |

### Institution names analysis

Among 26 unique studies that reported institution names, we compared the names with OHDSI collaborators (https://www.ohdsi.org/who-we-are/collaborators/). In total, we identified 92 unique institutions. Among them, 36 (38%) institutions’ names were found on the collaborator list. As suggested by Figure x1, Ajou University, Hanyang University, and Columbia University are the top contributors for the in-network sites. Seoul National University, Hallym University, and Sungkyunkwan University are the top contributors for the out-network sites.

**Appendix Figure 1.**
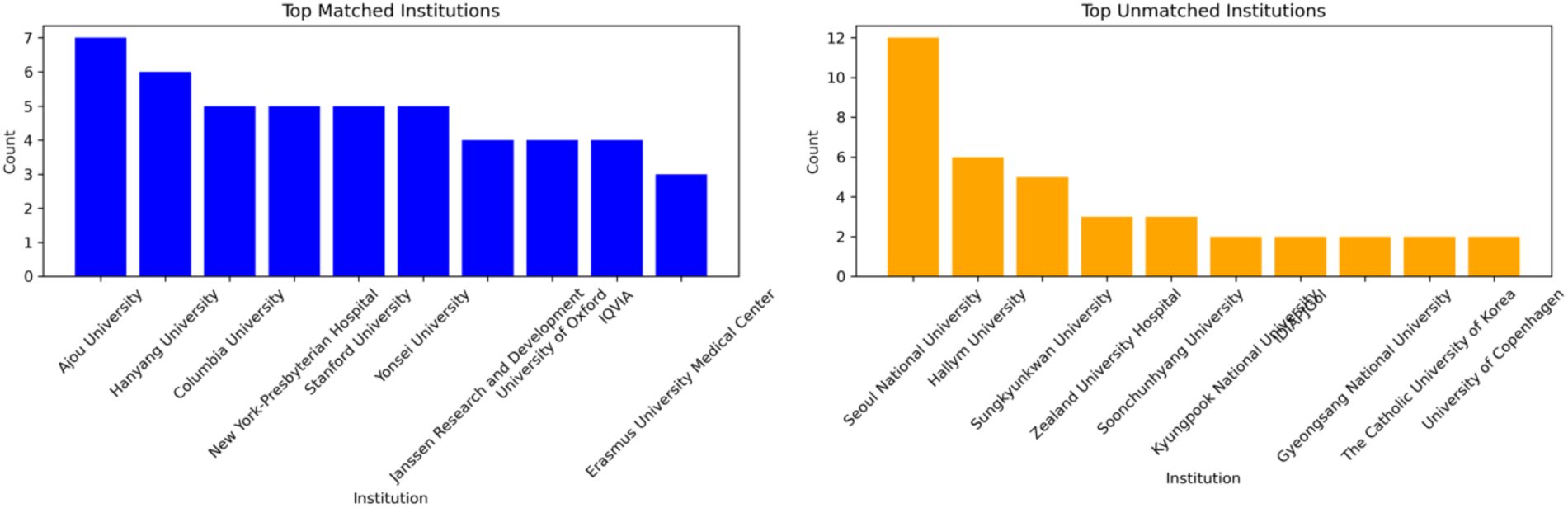
Distribution comparison between in-network (matched) and out-network sites.

**Appendix Figure 2.**
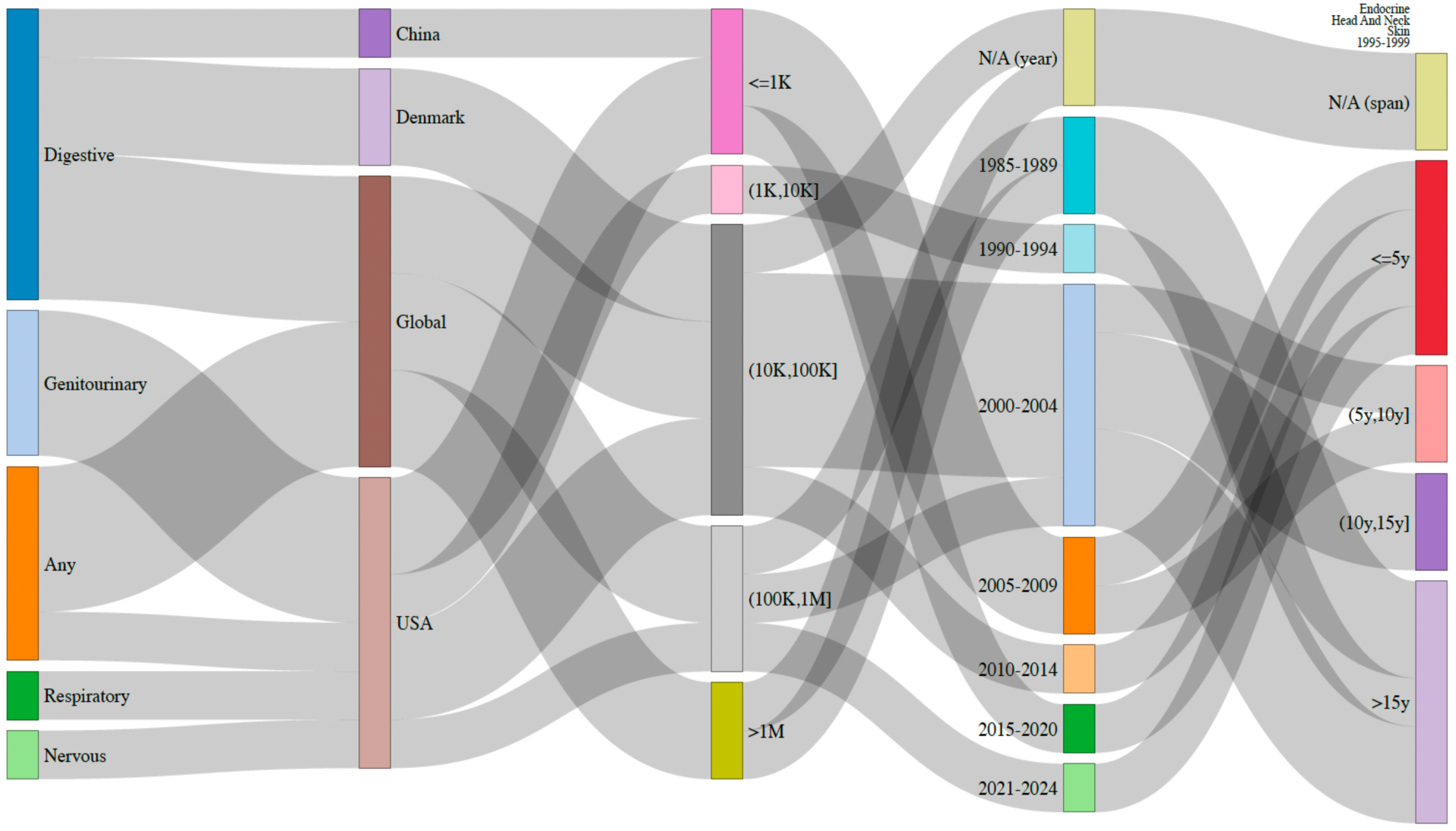
Linkage between the aggregated cancer type, geographic area, cohort size, start year of study, and study period. Analysis based on countries excluding South Korea.

### Appendix 1. Search Strategies

#### IEEE Xplore

27 articles resulted from: (cancer* OR tumor* OR tumour* OR neoplas* OR oncology*) in Abstract AND ("ohdsi"OR ("observational health data sciences Informatics") OR "omop" OR "observational medical outcomes partnership" OR "common data model") in Abstract AND (2010-2023 in Year)

#### Ovid

Journals@Ovid@TMC Library (subscribed full text)

Journals@Ovid (some full text)

Ovid MEDLINE(R) and Epub Ahead of Print, In-Process, In-Data-Review & Other Non-Indexed Citations and Daily <1946 to January 12, 2024>

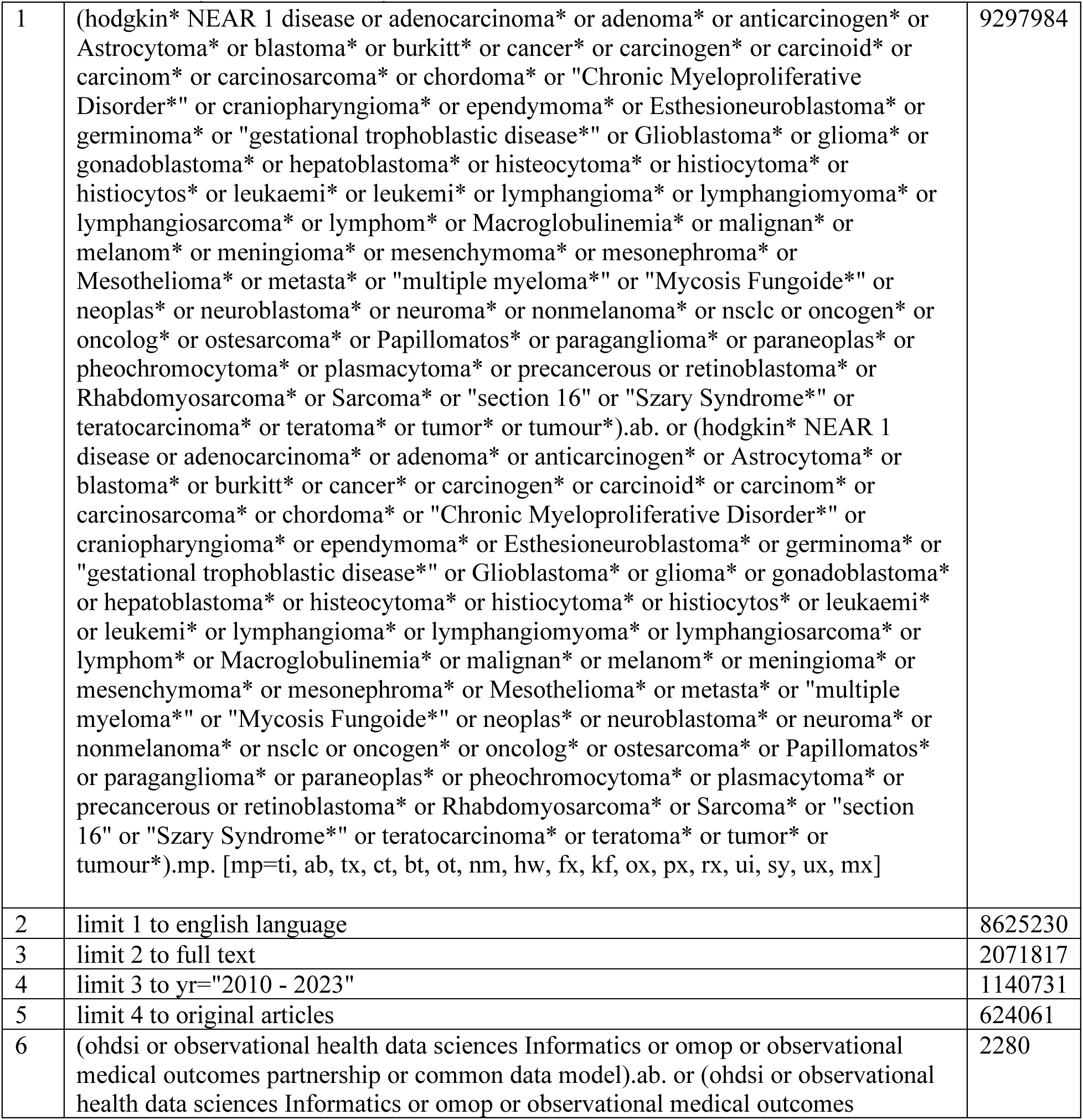
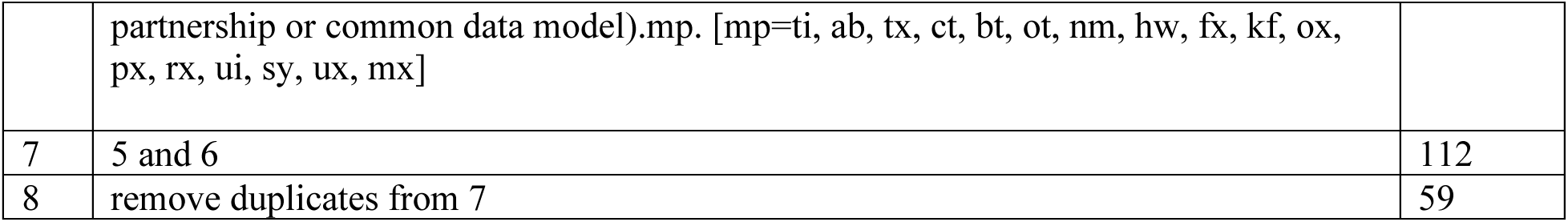

#### PubMed

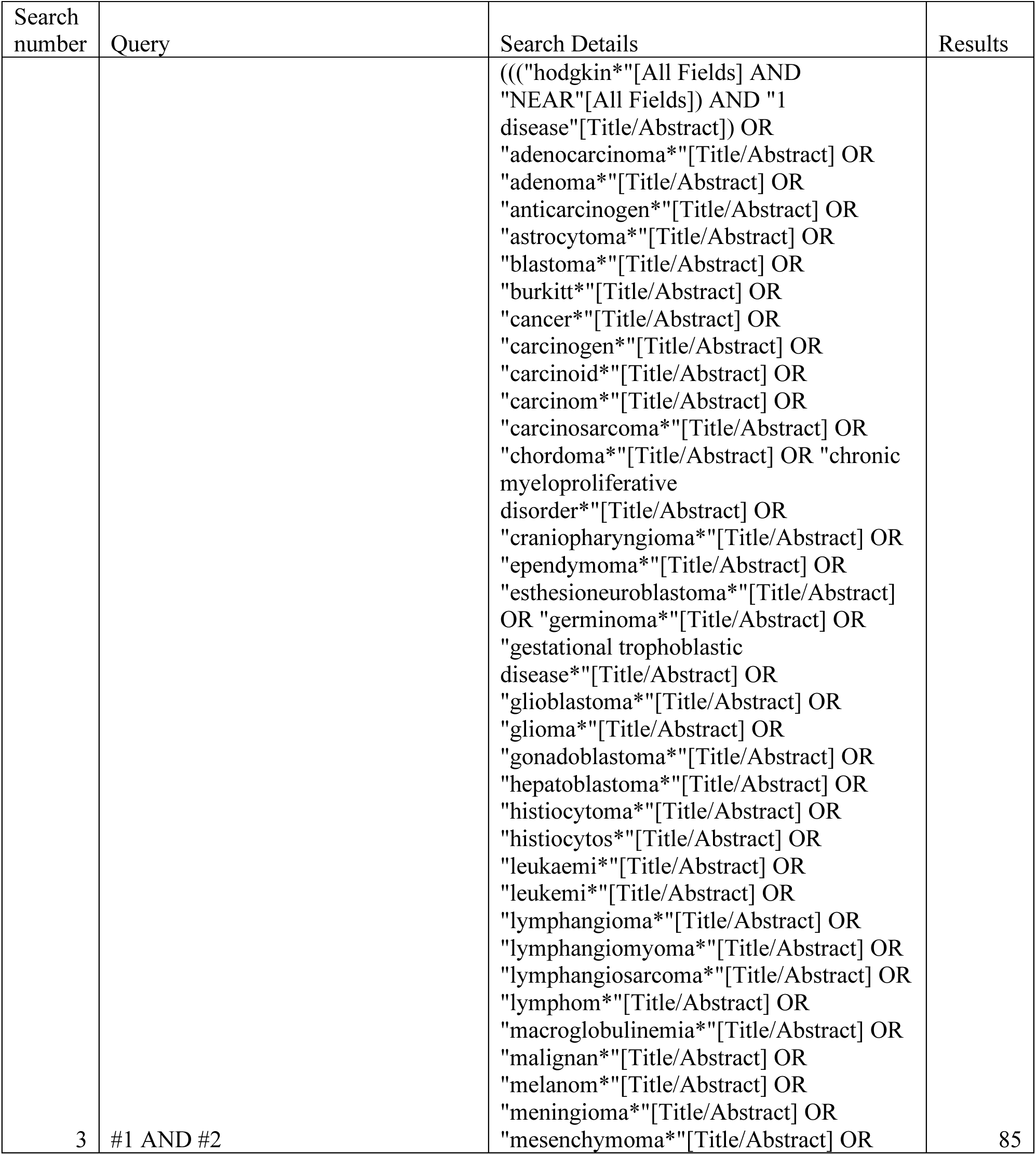
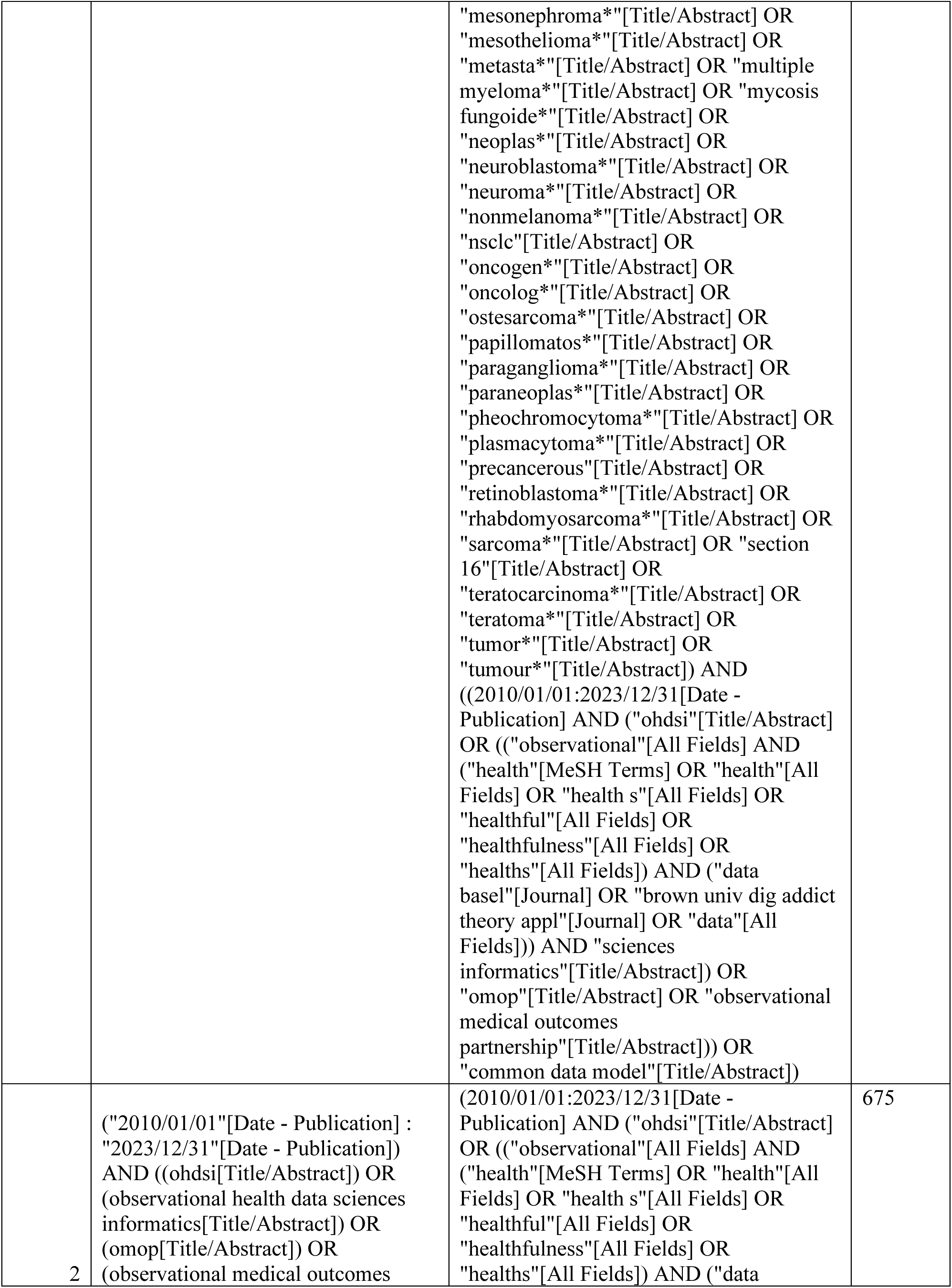
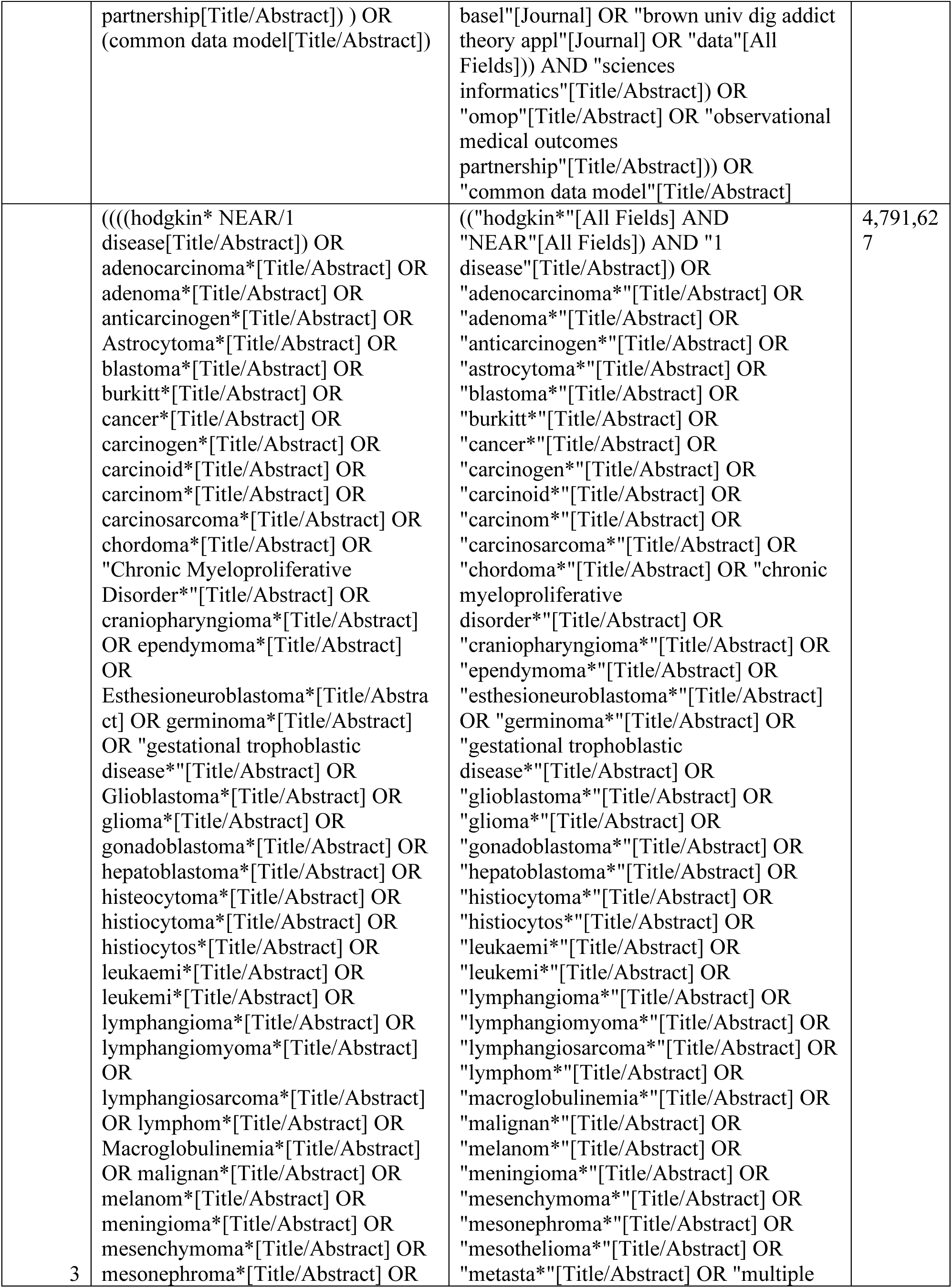
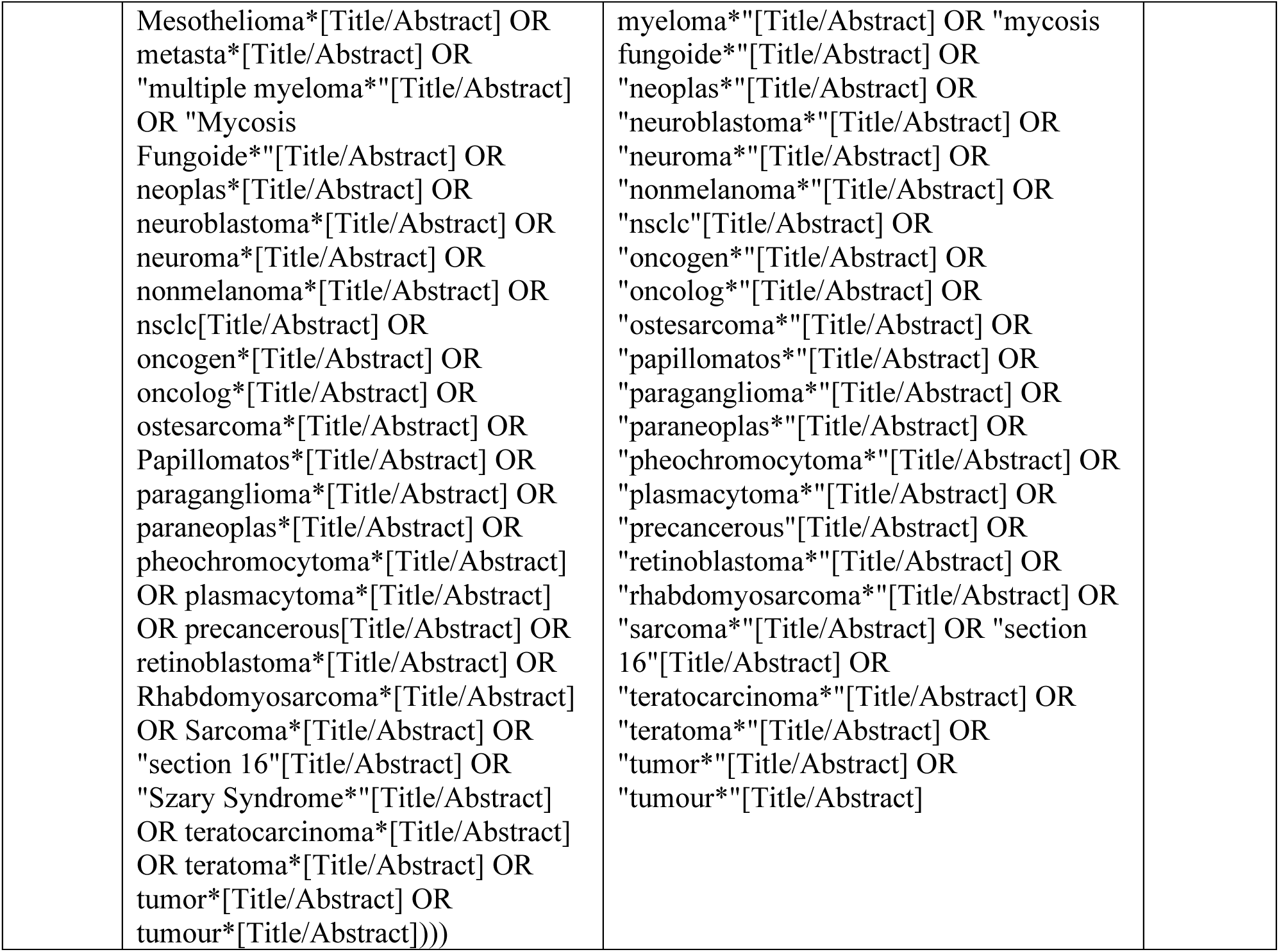

#### Web of Science

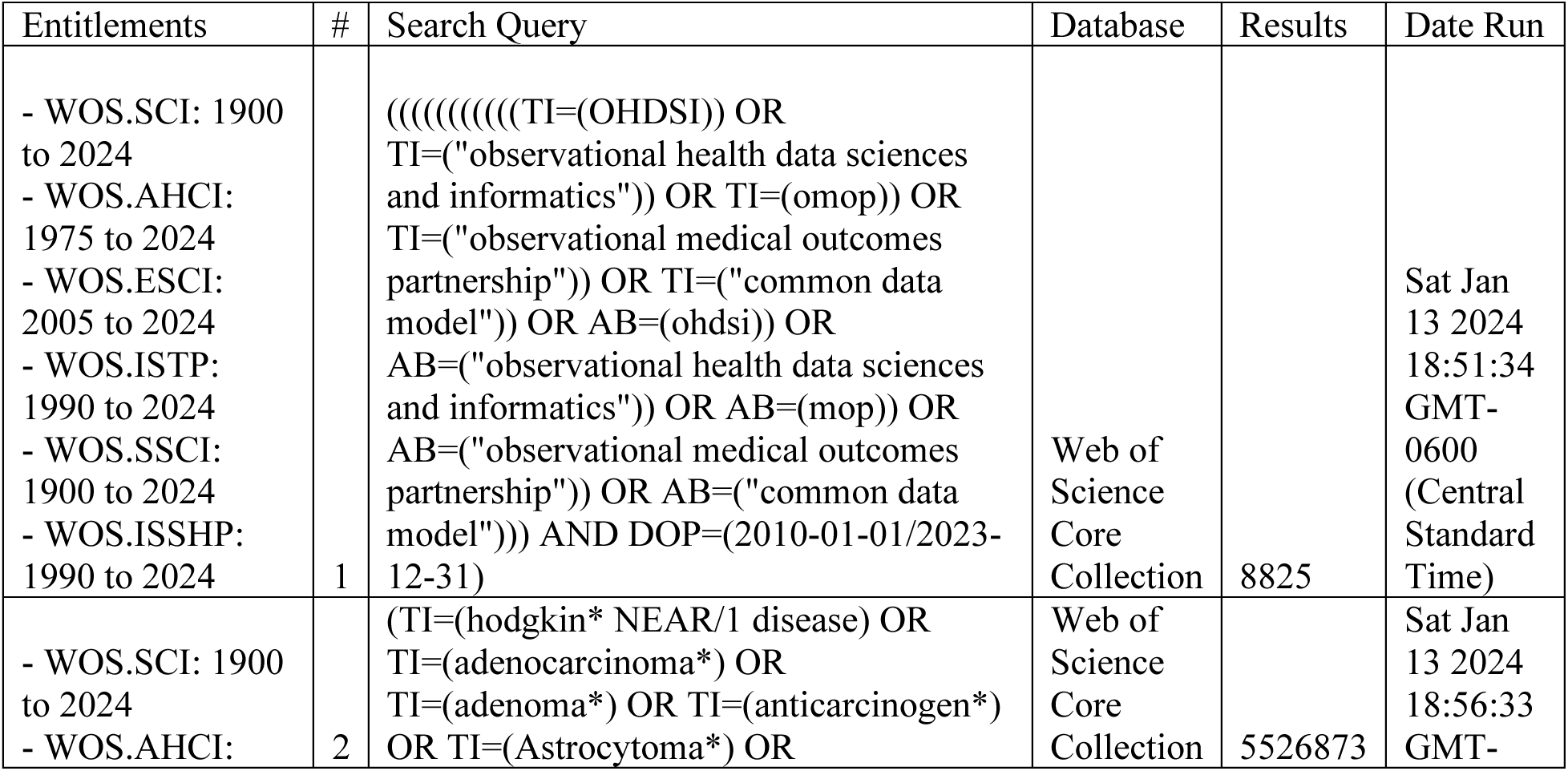
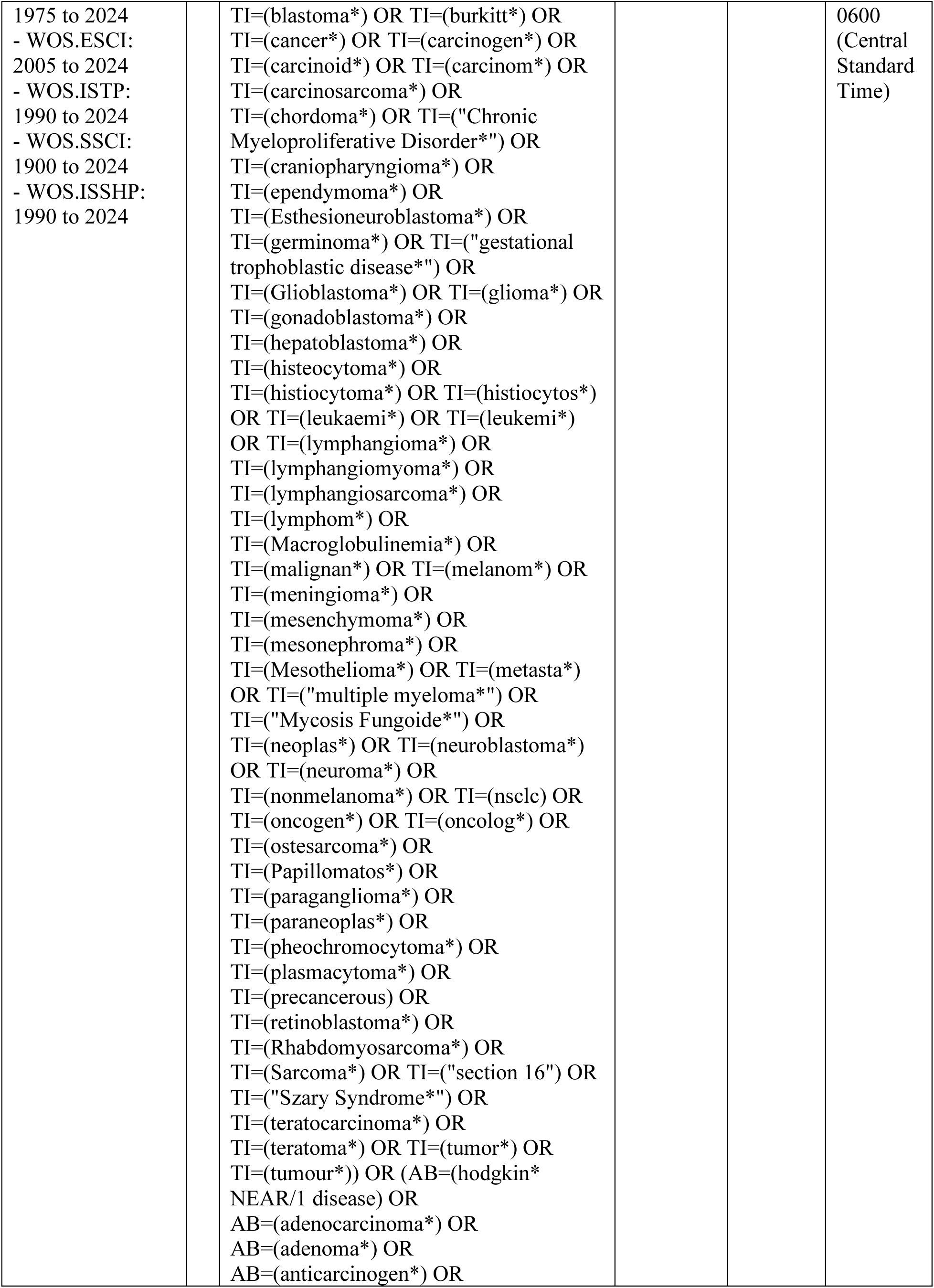
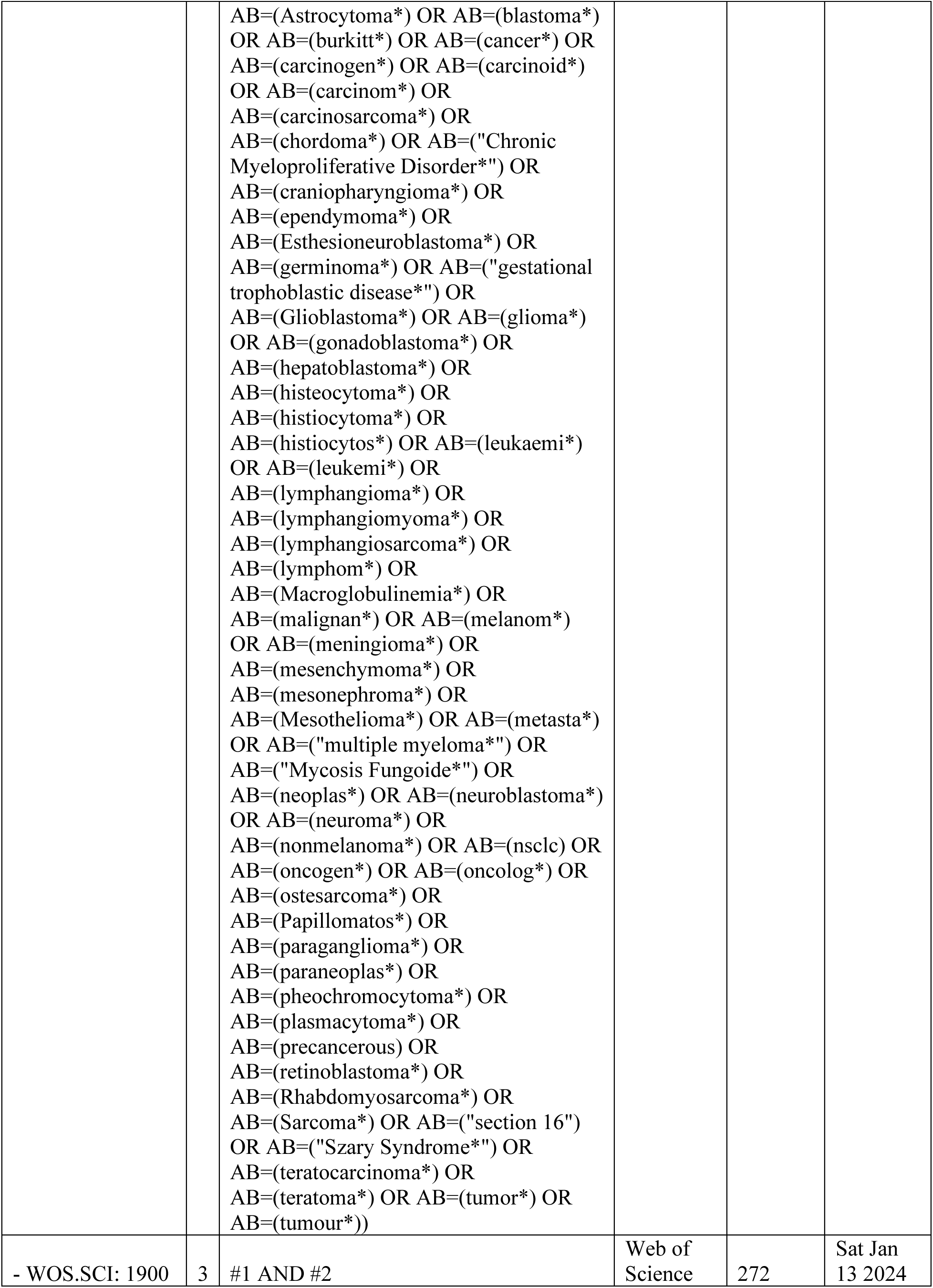
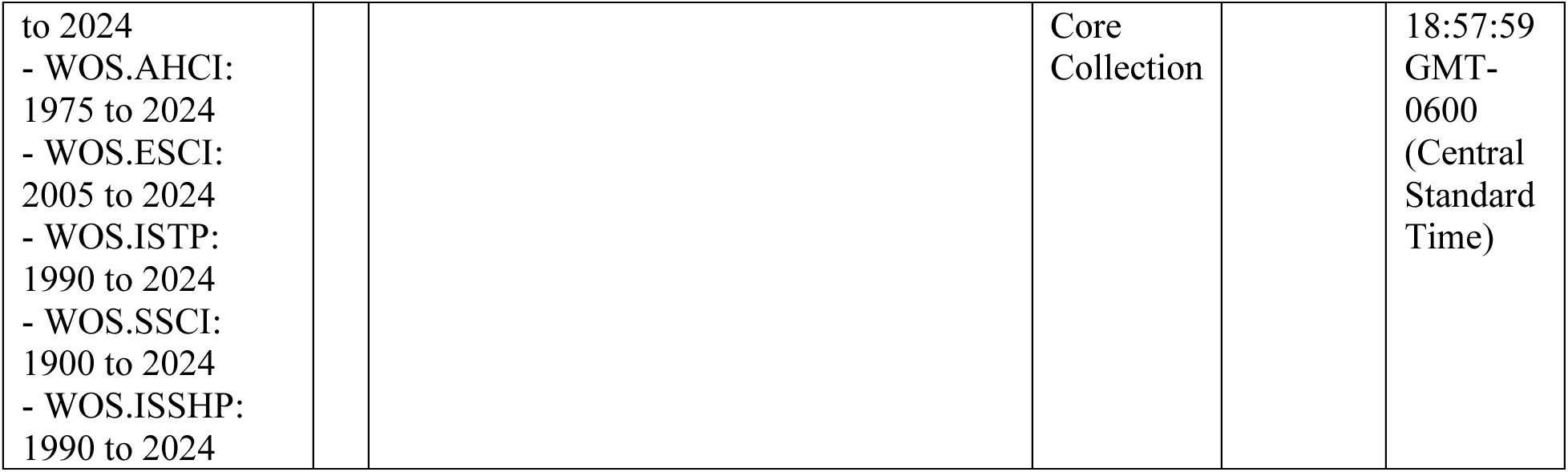

